# Third dose vaccine With BNT162b2 and its response on Long COVID after Breakthrough infections

**DOI:** 10.1101/2021.11.08.21266037

**Authors:** Ashraful Hoque, Md Marufur Rahman, Hossain Imam, Nurun Nahar, Forhad Uddin Hasan Chowdhury

**Affiliations:** Blood Transfusion, Sheikh Hasina National Institute of Burn & Plastic Surgery, Bangladesh; Department of Oncology and Metabolism, The Medical School, University of Sheffield, UK; Burn & Plastic Surgery, Sheikh Hasina National Institute of Burn & Plastic Surgery, Bangladesh; Dhaka Medical College Hospital,Bangladesh; Internal Medicine, Dhaka Medical College Hospital, Bangladesh

**Keywords:** Breakthrough infection, Long COVID, CRP

## Abstract

**Background:** Breakthrough events are not rare after emerging of Delta variant. On the other hand, long COVID is an unsolved issue where sufferers suffer a lot. Some study has shown that COVID-19 vaccine has improved some clinical and libratory parameters in long COVID. But what will be the possible measures against long COVID after the breakthrough event is still a burning question.

**Method:** We have observed the third dose by BNT162b2 in a small group(n=20) who were diagnosed as long COVID after breakthrough infections, in Sheikh Hasina National Institute of Burn & Plastic Surgery Institute, Dhaka, Bangladesh. CRP(C-reactive protein) and Anti S1 RBD IgG responses were measured.

**Result:** All 20 participants in the study received both dosage of “ChAdOx1-nCoV-19” in between February 2021 to April 2021 and had breakthrough infection in the same or following month which led to long COVID syndrome. They all received a third dose of “BNT162b2”. A before and after 3^rd^ dose (14 days after) CRP from participants serum was measured. A Wilcoxon matched paired signed rank test revealed significant (P value <0.05) reduction of inflammatory marker (CRP) after receiving the 3^rd^ vaccine dose. Pre and post 3^rd^ dose quantitative anti S1-RBD IgG response was measured and compared that revealed significant boosting effect that clearly correlates with the CRP response.

**Conclusion:** Coverage of vaccines all over the world is still not expected level to control this pandemic. WHO has not recommended the use of a third/booster dose of COVID vaccines. Though our results show some sort of hope for the long COVID in breakthrough events after getting the third dose more study is needed to conclude this issue.

## Introduction

From the beginning of COVID-19 spreads around the globe, it is mutating, which means it is acquiring genetic changes. This change has made vaccines difficult to combat and control the pandemic scenario. In the trial results of the vaccine, we have found that it can reduce hospital admission to a great extent. Now data showing on October 21, 2021, 35% of the 519 patients hospitalized with COVID-19 in Massachusetts had been fully vaccinated^1^. Multiple reports have documented that fully vaccinated individuals have become infected; their viral loads may be as high as the levels seen in unvaccinated individuals^2^. Patients with COVID-19 mostly return to baseline after acute infection, but some continue health issues. Throughout the world, what amount of people have been affected by long COVID after acute COVID remains unknown, but published data indicate that about 10-20% of COVID-19 patients suffer persisting symptoms for weeks to months following acute COVID-19 infection.

According to the Delphi consensus published on 6th October of this year, a post-COVID-19 condition usually occurs in individuals who had a history of probable or confirmed COVID-19 infection, usually 3 months from the onset of COVID-19 with symptoms that last for at least 2 months and cannot be explained by an alternative diagnosis. Brain fog, cognitive dysfunction, shortness of breath, is common presentations. Symptoms may be new-onset following initial recovery from an acute COVID-19 episode or persist from the initial illness. Symptoms may also fluctuate or relapse over time^3^.

CDC’s has defined a breakthrough infection as a COVID case that occurs in someone who is fully vaccinated, meaning 14 or more days after completing the recommended doses of an authorized vaccine.

No specific protocol is yet to be found to treat long COVID. Pharmacological management is still not good enough to cure it. We want to evaluate the role of the third dose of COVID vaccine in patients suffering from long COVID after breakthrough infection.

## Method

This observational study was conducted at Sheikh Hasina National Institute of Burn and Plastic Surgery, Dhaka February 2021 to October 2021. We enrolled 20 participants for our study who received both doses of ChAdOx1-nCoV-19 of the vaccine in the initial rollout of a vaccine in the country (February to April) and had breakthrough infection in the same or following month which led to long COVID syndrome. Participants’ demographic data along with past SARS-COV-2 infection history and existing comorbidities were recorded using a structured questionnaire. Serological testing for CRP was done by Invitrogen CRP Human ELISA kit by ThermoFisher Scientific and antibodies to the RBD of the S1 subunit of the viral spike protein (IgG) was performed from the plasma samples using a commercially available Anti-SARS-CoV-2 ELISA kit(Diasino, China) as per instruction supplied by the manufacturer after getting the third dose by BNT162b2. The study protocol was reviewed and approved by the Institutional Review Committee of the Sheikh Hasina National Institute of Burnand Plastic Surgery, Dhaka (approval memo number: SHNIBPS/July-14(09)). Informed written consent was taken from each participant. Data were summarized by descriptive analysis. Numerical variables are reported as mean and standard deviation or median and interquartile ranges, as appropriate. Categorical variables are reported as counts and percentages. The analysis was performed in Graphpad Prism 9.0.

## Result

All 20 participants in the study received both dosage of “ChAdOx1-nCoV-19” in between February 2021 to April 2021 and had breakthrough infection in the same or following month which led to long COVID syndrome. The most commonly reported symptoms were unusual fatigue, palpitation and insomnia. They all received a third dose of “BNT162b2”. A before and after 3^rd^ dose (14 days after) CRP from participants serum was measured. A Wilcoxon matched paired signed rank test revealed significant (P value <0.05) reduction of inflammatory marker (CRP) after receiving the 3^rd^ vaccine dose (Figure 1).

**Figure 1:**
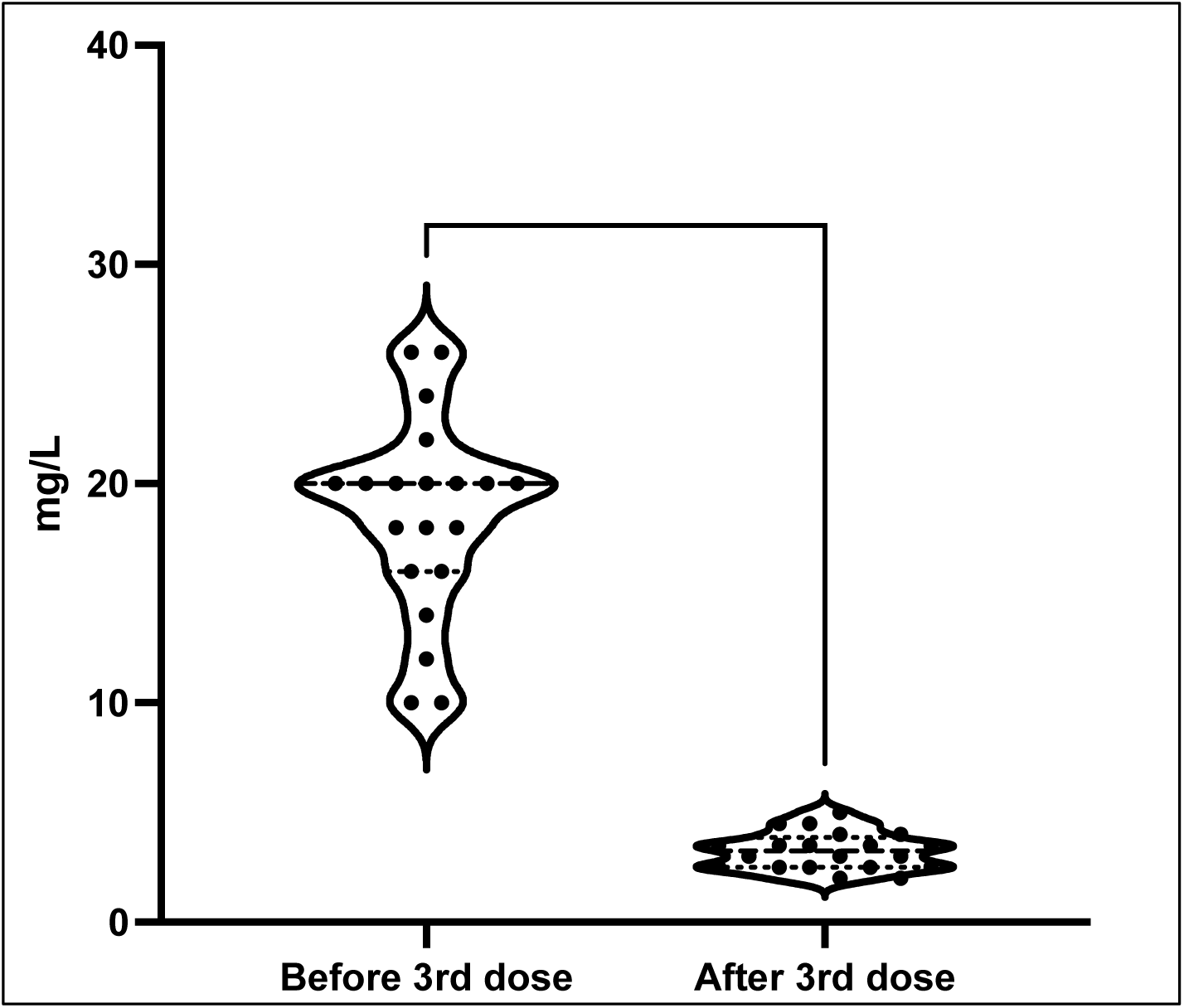
Comparison of serum CRP level before and after receiving 3^rd^ dose of COVID-19 vaccine in individuals with long covid syndrome

Pre and post 3^rd^ dose quantitative anti S1-RBD IgG response was measured and compared that revealed significant boosting effect (Figure 2) that clearly correlates with the CRP response shown in Figure 1.

**Figure 2:**
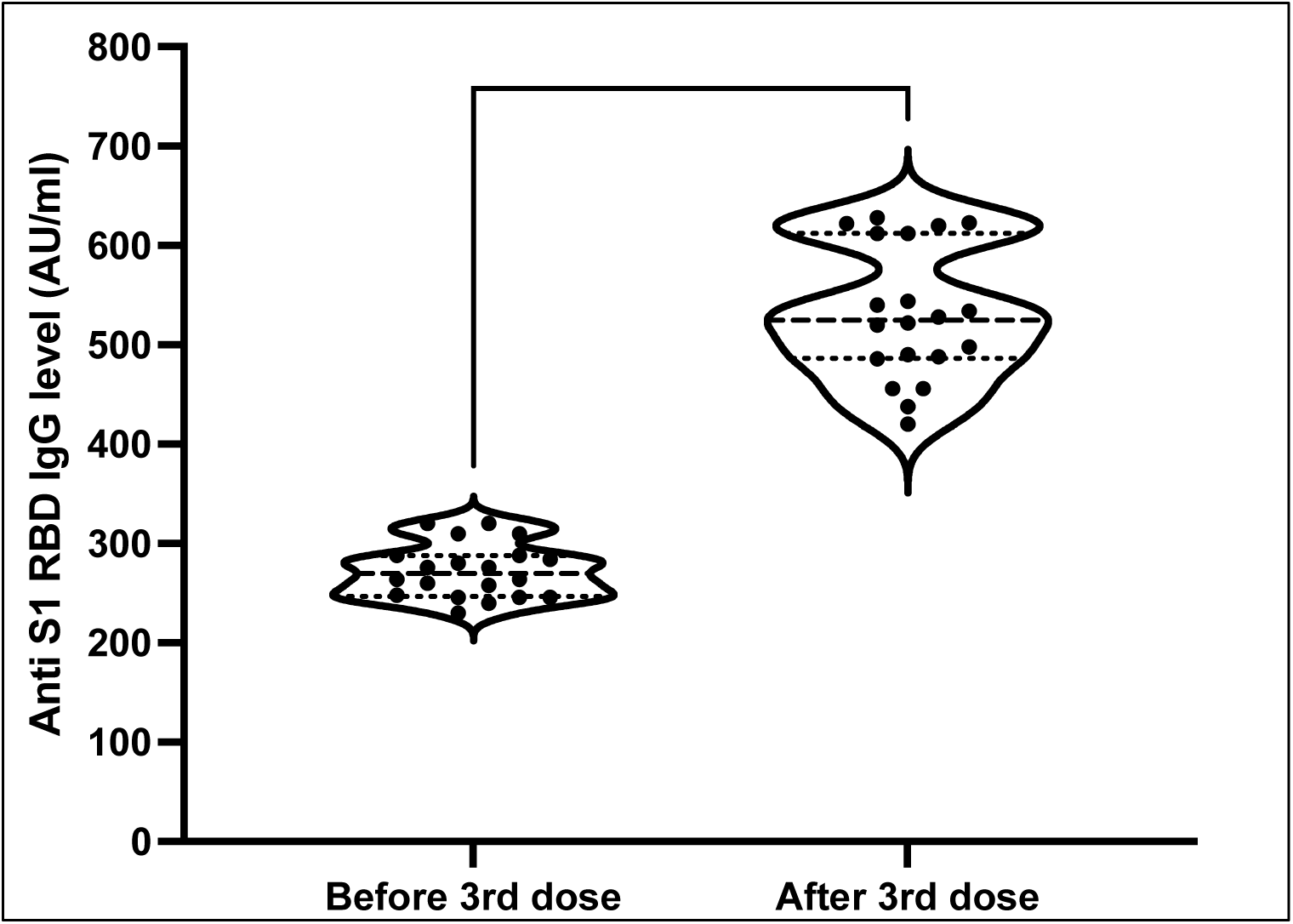
SARS-CoV-2 Anti S1 RBD IgG comparison before and after receiving a 3^rd^ dose of COVID-19 vaccine.

## Discussion

Most COVID-19 vaccines are effective at preventing most infections. But no vaccine is 100% is effective. So breakthrough infections are expected.

On the other hand, long COVID may be driven by long-term tissue damage like brain, lungs, heart, etc caused by inflammation such as autoimmunity, immune dysregulation viral persistence. Female sex, early dyspnoea, prior psychiatric disorders, and specific biomarkers (e.g. D-dimer, CRP, and lymphocyte count) may associate risk factors may include. More research is needed to establish such risk factors^4^.

One known aspect of long COVID is that similar post viral syndrome was observed with prior human corona virus diseases. For example, symptoms of fatigue, myalgia, and psychiatric impairments have inflicted survivors of Middle East respiratory syndrome (MERS) and severe acute respiratory syndrome (SARS) for up to four years ^5-9^.

Even at 7-year and 15-year follow-ups, pulmonary and bone radiological complications were still evident among a proportion of SARS survivors who were mostly younger than 40 years^10,11^. A review has proposed that T-cells dysfunction may promote long COVID pathophysiology similarly in autoimmune diseases^12^.

SARS-CoV-2 could make antigen-presenting cells present antigens to auto-reactive T-cells in a process called bystander activation. This is consistent with autopsy examinations of deceased COVID-19 patients showing that infiltrates in the lungs and other organs were enriched with CD8þ T cells, one of the crucial mediators of autoimmune reactions^13^.

B-cells may also be involved in long COVID autoimmunity. In a study analysing serum samples from hospitalized COVID-19 patients, antiphospholipid, autoantibodies were detected in 52% of samples, which were further associated with neutrophil hyperactivity and more severe clinical outcomes^14^. Besides, evidence exists that severe COVID-19 causes lymphopenia (i.e. B-cell and T-cell lymphocytes deficiency) that causes hyperinflammation^15,16^. This is because lymphocytes, particularly T-cells, participate in inflammation resolution following infection^17,18^. Meta-analyses have determined lymphopenia and high pro-inflammatory neutrophil count as independent risk factors of COVID-19 severity and mortality^19-21^. Thus, as B-cell and T-cell lymphocytes are renewed, elevated inflammation from unresolved hyper inflammation may ensue and contribute to longCOVID^22-24^.

Moreover, decreased T-cell and B-cell numbers have been shown to correlate with persistent SARS-CoV-2 shedding, which may further perpetuate chronic immune activation in long COVID^25,26^. Residual inflammation from post-SARS-CoV-2 could lead to long COVID in adults. Raised pro-inflammatory markers (e.g. CRP, IL-6, and D-dimer) and lymphopenia have been associated with long COVID.

Other studies have shown that COVID-19 pulmonary lesions at two-month post-admission were associated with elevated systemic inflammatory biomarkers, such as D-dimer, interleukin-6 (IL-6), and CRP^27,28^.

A study showed, increased D-dimer and CRP levels and decreased lymphocytes were more common in COVID-19 survivors who developed persistent symptoms than their fully recovered counterparts^29^.

Response to infections, liver synthesizes significant quantities of acute-phase proteins (APPs), such as CRP ^30,31^. Acute inflammatory protein is a highly sensitive biomarker for inflammation, tissue damage, and infection ^32^. Different result has shown that CRP levels are correlated with levels of inflammation ^33^. CRP levels can promote phagocytosis and activate the complement system ^34^. In other words, CRP binds to microorganisms and promotes their removal through phagocytosis ^35^.

Long COVID SOS a survey conducted by UK advocacy group, the University of Exeter, and the University of Kent which showed 57% of participants experienced at least some improvement in symptoms after a vaccine when they were affected by long COVID. Less than 7% have experienced deteriorating symptoms. Though all vaccines showed benefits they have said mRNA vaccine improved the scenario most.

One of the possible hypotheses behind the improvement after getting the vaccine is, persistent infection causes long-haulers to feel sick, and then the vaccine shot might be helping their immune system finally clear the virus. Another possible theory is that vaccines might rev up a person’s immune system and shift the sufferers back into a steady state of equilibrium.

In our study, we have found a dramatic improvement in CRP value which was higher than normal after getting COVID several months back. Their Ct values of rt-PCR were persistently around 36. These changes can be a possibility of immune boost up after the third dose of vaccine which initiates viral clearance from the body. Quantitative assessments of antibodies (IgG) were raised after getting the third dose.

According to these two results with their improvement of symptoms, we hypothesize that the third dose with BNT162b2 might clear the residual viral load as it could halt the ongoing inflammatory process initiated by breakthrough infections.

### Limitations and Future Suggestions

This short sample size cannot produce generalizability. Therefore, subsequent clinical studies with larger sample sizes with different age group should be enclosed to confirm our findings.

## Conclusions

The third dose is still not recommended by most the country in the world as well as WHO as vaccines are not sufficient according to the demand of pandemics. Few countries like the USA approved the third dose to combat against emerging Delta variant. Long COVID is a very notorious condition that has no proper solution to date. From our study, we can hope that the third dose with mRNA vaccine could be an option to control it. More data is needed to conclude this issue.

## Data Availability

All data produced in the present work are contained in the manuscript

